# Isoniazid preventive therapy completion and factors associated with non-completion among patients on antiretroviral therapy at Kisenyi Health Centre IV, Kampala, Uganda

**DOI:** 10.1101/2022.11.03.22281894

**Authors:** Ian Amanya, Michael Muhoozi, Dickson Aruhomukama, Anthony Ssebagereka, Richard Mugambe

## Abstract

**Background:** Isoniazid preventive therapy (IPT) is given to HIV patients to reduce the risk of active tuberculosis (TB). However, treatment completion remains sub-optimal among those that are initiated. This study aimed to determine the completion level of IPT and the factors associated with non-completion among people on antiretroviral therapy (ART) at Kisenyi Health Center IV in Kampala, Uganda.

**Methods:** A facility-based retrospective cohort study utilizing routinely collected data of 341 randomly selected HIV patients initiated on IPT was conducted. Data extracted from the registers were used to determine the IPT completion. Modified Poisson regression with robust error variances was used to determine the associated factors of IPT non-completion while in-depth interviews were conducted to explore barriers to IPT completion from the patient’s perspective.

**Results:** A total of 341 patients who started on isoniazid (INH) were retrospectively followed up, with 69% (236/341) being female. Overall IPT completion was at 83%. Multivariable analysis revealed the prevalence of IPT non-completion among males was 2.24 times the prevalence among females [aPR 2.24, 95% CI: 1.40-3.58]. The prevalence of IPT non-completion among patients with a non-suppressed HIV viral load was 3.00 times the prevalence among those with a suppressed HIV viral load [aPR 3.00, 95% CI: 1.44-6.65]. Patients who were married/cohabiting had a 69% lower prevalence of IPT non-completion compared to those who were single [aPR 0.31, 95% CI: 0.17-0.55]. Lack of IPT-related health education, pill burden, distance to the health facility, and patient relocation were reported as the barriers to IPT completion.

**Conclusion:** IPT completion was found to be at 83% among the cohort studied. However, lower completion levels persist among males and HIV virally non-suppressed patients. Lack of IPT-related health education, pill burden, distance to the health facility, and patient relocation were reported as the barriers to IPT completion. Interventions that target these groups of people need to be intensified.

## Background

The World Health Organization (WHO) estimates a quarter of the world’s population to be infected with latent Tuberculosis of which the majority live in sub-Saharan Africa which is home to about 14% of the global population (1). An estimated 10 million people developed TB disease in 2017 of whom 72% were living in sub-Saharan Africa, of those that developed TB disease, 9% were co-infected with HIV (1). Immune suppression particularly due to HIV infection greatly increases the risk of TB infection as well as latent TB progression to an active infection (2, 3). The risk of developing TB is 20 to 37 times greater in people living with HIV (PLHIV) than among those who do not have HIV (4). Antiretroviral therapy (ART) has been reported to reduce the risk of acquiring TB (5) though the risk remains higher in people with HIV; and these people are up to four times more likely to die during TB treatment (6, 7).

Uganda had an estimated 1.4 million people living with HIV in 2018, of whom 72% were on ART (8). Uganda also registered about 86,000 new TB infections in 2017 of whom an estimated 40% were co-infected with HIV. Among these HIV patients, TB accounted for about 30% of all deaths (1). In Kampala, the prevalence of HIV is estimated to be about 6.9%, which is higher than the national average of 6.2% (9). Additionally, the Uganda national TB prevalence survey of 2014/15 revealed a higher bacteriologically confirmed (B+) TB prevalence of 504/100,000 in urban areas compared to 370/100,000 in rural areas (10). Kampala had the highest TB case notifications in Uganda suggesting that TB was more common in urban areas than in rural areas.

WHO recommends the use of three I’s (Isoniazid Preventive Therapy (IPT), intensified case finding, and infection control) to reduce the risk, and burden of TB in people living with HIV (11). IPT use reduces the risk of TB disease among HIV-infected individuals by about 60% (12, 13), and this efficacy is even higher accompanied by ART (14). IPT also prevents the progression of latent TB to active TB disease (15).

In 2014, Uganda adopted WHO’s guidelines that recommended the use of IPT for TB prevention as part of the comprehensive HIV/AIDS care strategy (16). Despite the available evidence that IPT is effective, many countries including Uganda have had little success in implementing this recommendation (1). In Uganda, the uptake of IPT as part of the TB prevention strategy remains low and this has mainly been attributed to limited funding for IPT commodities, lack of awareness of the potential benefits of IPT, health facilities, district & implementing partners not being held accountable for IPT performance; lack of data utilization at national, district & health facility level and lack of sustained quality mentorship among other factors (17).

In addition, adherence and completion of treatment among patients initiated on IPT remain major concerns as these affect its effectiveness (14). Limited studies conducted in Ethiopia, Uganda, and Malawi respectively, have documented completion levels of IPT ranging from 36 to 98% (18-20).

However, few studies had documented patient reasons for failure to complete the treatment in Uganda. Understanding this was key to improving the use of IPT among ART patients. Using routinely collected data, this study aimed to assess the completion of IPT and factors associated with non-completion among patients on ART. This would inform stakeholders, help to improve IPT use and help realize its benefits thereby reducing the risk and burden of TB among these people.

## Materials and methods

### Study area

Kampala, Uganda’s capital has an estimated resident population of 1.5 million people (21). However, the total population attracted by the various economic activities and services in almost doubles the resident population. The study was conducted at Kisenyi health centre IV in Kampala’s central division. Kisenyi health centre IV was purposively selected as it’s the largest operational health center IV in Kampala that also receives a high volume of patients in comparison to smaller health facilities that provide HIV care. The facility is financed by the government but also receives funding from donors through implementing partners. Services offered by this facility are free and include TB services, laboratory services, dental services, HIV counseling and testing, maternity services, Antenatal/EMTCT services, immunization, Early Infant Diagnosis (EID), Nutrition services, Family planning services, Postnatal services, Cancer screening, STI management, youth-friendly services, and comprehensive ART outreaches among others. The ART and TB clinics are integrated and open from Monday through Friday to accommodate the nearly 12000 active ART patients. To increase completion rates, IPT and ARV medicine refills are given out simultaneously with synchronized durations. A number of tools, including the ART dispensing logs, IPT registers, ART cards, the ART register, the viral load log, and the daily activity consumption log, among others, record the drugs that have been prescribed. The patient appointment and visit dates, patient ID, current ART regimen, number of drug days distributed, weight and height, and whether the patient was bled for viral load or CD4 are just a few of the details recorded in these instruments. Active registers are kept at the various service stations, and the tools are housed in a separate records room. The Uganda Electronic Records Management system (UgandaEMR), which is located in the records sector, also stores this data electronically.

### Study design

This was a facility-based retrospective cohort study utilizing routinely collected data. Quantitative data extracted from the facility registers were used to answer the questions on completion of IPT and the factors associated with non-completion while qualitative data collected through in-depth interviews with patients were used to explore the barriers to IPT completion from a patient’s perspective.

### Study population

The study participants were HIV patients on ART at Kisenyi HC IV that had been initiated on IPT. To reduce recall bias, ART patients that had been initiated on IPT in September 2019 at Kisenyi HC IV were considered.

### Inclusion criteria

Only ART patients initiated on IPT in September 2019. Purposively selected non-completing ART patients for in-depth interviews who provided informed consent. Minors who provided assent and whose caretakers provided consent were included in the study.

### Exclusion criteria

ART patients initiated on IPT in any period other than September 2019 and those whose records could not be traced or found at the facility were excluded from the study. Patients who were ill, unavailable at the time of data collection and those who had no phone contacts were excluded from in-depth interviews.

### Sample size considerations

To determine the completion of IPT and the factors associated with non-completion, the Yamane formula at a 0.05 level of precision was used (22). This formula was used due to the finite population of patients initiated on IPT.

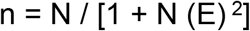

Where; n = sample size, N = population size, E = level of precision.

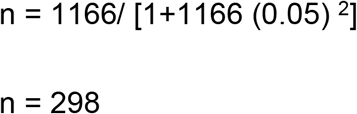

Adjusting for a 10% loss to follow-up

The minimum sample size required was 331.

### Sampling procedure

A list of the patient IDs initiated on IPT during September 2019, was compiled. A random sample of 341 patients was drawn, and their records were extracted from the IPT registration using Microsoft Excel. Respondents for in-depth interviews were purposively selected from the list of non-completers. The respondent selection was stratified by age group, sex, marital status, and district of residence (Table 1).

**Table 2:** Planned and conducted In-depth interviews.

|  | Planned Interviews | Conducted Interviews |
| --- | --- | --- |
| Characteristic | Number | Number |
| **Sex** |  |  |
| Male | 9 | 7 |
| Female | 9 | 8 |
| **Age group** |  |  |
| 0-9 | 3 | 0 |
| 10-19 | 3 | 1 |
| 20-29 | 3 | 5 |
| 30-39 | 3 | 4 |
| 40-49 | 3 | 3 |
| 50+ | 3 | 2 |
| **Marital status** |  |  |
| Single | 5 | 4 |
| Married/cohabiting | 5 | 6 |
| Separated/divorced | 5 | 5 |
| Widowed | 3 | 0 |
| **District of Residence** |  |  |
| Kampala | 6 | 8 |
| Wakiso | 6 | 6 |
| Others | 6 | 1 |

In a phone call, the participants gave their permission to participate in the form of audio that was recorded. Interviews continued until they reached saturation.

### Study variables

#### Dependent variable

The IPT non-completion, which was measured as a person failing to pick up their fifth consecutive monthly refill within 6 months of initiation, was the dependent variable.

Patients who had begun IPT had to return to the medical facility every month for five months starting after a 2-week assessment for any adverse effects. IPT refills were given by clinicians at the monthly visit, and they also evaluated the patient’s adherence and side effects. INH was prescribed for multiple months to patients who received ART on a multi-month dispensation.

It is important to note that the Ministry of health recommends that IPT should be given to all patients with the following criteria: HIV-positive children (≥one year of age), adolescents, and adults with no signs and symptoms of TB; HIV-positive infants and children <5years who have no signs and symptoms and active TB disease who get in contact with a person with active TB disease, irrespective of previous TPT; HIV-positive pregnant mothers with a history of contact with a TB patient after ruling out active TB; HIV-positive pregnant mothers with a WHO Stage 3 or 4 event and/or CD4 <200 without active TB (23).

#### Independent variables

The independent variables included; age, sex, Body Mass Index (BMI), marital status, WHO HIV clinical stage at IPT initiation, and pre-IPT HIV viral suppression (defined as the most recent measurement within 12 months to IPT initiation).

### Data collection tools

The qualitative data was collected using a call recorder and an in-depth interview guide, and the quantitative data was extracted using a data extraction tool.

### Data collection procedures

Data collection ran for one month. Quantitative data were extracted from the IPT register, ART cards, and the Uganda Electronic Medical Records system (EMR) using a structured data tool. Variables captured quantitatively were; Residence (District, sub-county, village), WHO HIV stage at the time of IPT initiation, IPT refill and next appointment dates, weight, height, age, sex, pre-IPT viral load result, and marital status. In-depth interviews that lasted 20-30 minutes were conducted by a trained research assistant with the help of an interview guide. Variables explored qualitatively included; IPT-related health education, challenges encountered while on IPT, and smoking and/or alcohol consumption. These variables gave rise to some of the themes presented in the findings. The interviewer introduced herself to the respondent, informed him/her about the aim of the study, duration of the interview, potential risks or benefits of participating in the study, and then sought consent to take part in the study. All qualitative interviews were recorded with a call recording application and notes taken during the interviews.

### Quality control

#### Training of research assistants

Two graduate research assistants were recruited and trained to take part in the study to standardize data collection procedures. The training involved a review of the study objectives, variables to be extracted, data sources and collection procedures, ethics, and confidentiality. The training ensured that the research assistants were confident, knowledgeable, and comfortable with the extraction tools, data sources, and procedures.

#### Pre-testing of the tools

Both the data extraction tool and interview guide were pre-tested at Kawaala Health Centre III which is a public health facility in Rubaga division, Kampala district. The tools were then appropriately adjusted based on the results from the pre-test to ensure that all the available and relevant data was captured.

#### Field editing of data

The principal investigator supervised the data collection exercise regularly and met with the research assistants to address any issues that arose, share experiences, lessons learned, and guide the way forward. Accuracy and completeness of all filled data collection tools were emphasized and checked daily before submission.

### Data management and analysis

#### Data storage and entry

Raw data in form of complete tools were safely kept in a lockable shelf. The data was then entered into a password-protected data entry screen in MS Excel that was developed by the PI before being exported to STATA version 14.0 for cleaning and analysis. Recordings of in-depth interviews with unique identifiers were secured with a password, simultaneously transcribed, and translated to English verbatim. Each transcript was proofread by the PI for content accuracy and completeness before being exported to Atlas ti. version 6.0 for analysis. Final transcripts and dataset were securely stored on a password-protected computer and backed up on a secure cloud-based account.

### Data analysis

#### Quantitative data

**Data entry, cleaning, and sorting;** data was entered into a developed excel data screen, and checked for completeness and logical flow. The goal was to understand and identify variables to be used in the analysis. Output was a working dataset.

**Exploratory data analysis;** the nature of the variables was examined, new categories and variables were generated, and unwanted variables were dropped at this stage. The goal was to gain a deeper understanding of the data and variables. Output was an analytic dataset.

**Descriptive analysis;** Characteristics of study participants were described using frequencies categorical variables, mean, and SD for continuous variables. IPT non-completion was determined as a proportion that failed to pick up their 5^th^ consecutive monthly refill within 6 months among those initiated.

**Bivariable analysis;** simple modified Poisson regressions of the outcome and the predictor were run to determine factors associated with non-completion to be included in the multivariable model. The output was crude prevalence ratios and confidence intervals. Variables with a p-value <0.2 were selected for inclusion in the multivariable model.

**Multivariable analysis;** a modified Poisson regression model with robust error variances was run to determine the predictors of IPT non-completion. The output was adjusted prevalence ratios with corresponding confidence intervals. Variables were checked for multicollinearity before being added to the model. Variables with a p-value <0.05 were significantly associated with IPT non-completion.

#### Qualitative data analysis

All audio recordings were concurrently transcribed and translated to English verbatim. Transcripts were then proofread for accuracy by the PI before being uploaded to Atlas.ti Version 6.0 for analysis. Data analysis was conducted by a trained data analyst. Thematic analysis was used to analyze the data. Both inductive and deductive coding approaches were applied to generate the codebook (24, 25). The codebook generated during analysis was reviewed by the PI before adoption. After the review of codes with the help of the code families’ option in Atlas.ti, codes were then combined, others dropped or renamed to fit under emerging themes. The results were presented as narratives along with supporting quotes per theme. The analysis was regularly reviewed by the PI at every stage to ensure the reflexivity of the interpretations.

### Ethical considerations

Ethical approval for the study was sought and granted by Makerere University School of Public Health Higher Degrees, Research and Ethics Committee. Permissions were subsequently granted by the Director of Public Health Environment at KCCA and the In-charge Kisenyi HC IV. Recorded consent was also obtained from the respondents. The research assistants were trained on how to ensure privacy and confidentiality for all participants both during and after data collection. Data collection took place at the height of the COVID-19 Pandemic. As result, research assistants were provided with adequate masks and sanitizer as part of infection prevention and control (IPC) measures. In addition, In-depth interviews were conducted over the phone in place of physical interviews for IPC reasons. Consequently, participant consent was audio recorded.

## Results

### Description of the study population

A total of 341 records of participants were included in the analysis. About 31% (104/341) of the participants were aged 29 years and below. The median age was 33 (IQR 28 - 40). The majority of the respondents were female comprising 69% (236/341) and 54.5% (186) had a normal body weight. Additionally, 97.1% (331/341) of the participants were at the WHO HIV clinical stage1. All the participants had a viral load test done in the last 12 months with 97% (330) having a suppressed result. Regarding marital status, 58.4% (199/341) of the participants were married/cohabiting while 20.5% (70) were Divorced/separated (Table 3).

**Table 4:** Characteristics of the study population.

| Characteristic | Frequency |  |  |
| --- | --- | --- | --- |
| Age | Non-Completers %<br>(n)<br>17% (58) | Completers %<br>(n)<br>83% (283) | Overall<br>(n) |
| 0-29 | 20.2% (21) | 79.8% (83) | 104 |
| 30+ | 15.6% (37) | 84.4% (200) | 237 |
| **Sex** |  |  |  |
| Female | 13.6% (32) | 86.4% (204) | 236 |
| Male | 24.8% (26) | 75.2% (79) | 105 |
| **BMI** |  |  |  |
| Normal weight | 19.9% (37) | 80.1% (149) | 186 |
| Underweight | 22.2% (4) | 77.8% (14) | 18 |
| Pre-obesity | 14.1% (12) | 85.9% (73) | 85 |
| Class 1 obesity | 12.1% (4) | 87.9% (29) | 33 |
| Class 2&3 obesity | 5.3% (1) | 94.7% (18) | 19 |
| **WHO HIV clinical stage** |  |  |  |
| Stage 1 | 16.9% (56) | 83.1% (275) | 331 |
| Stage 2&3 | 20.0% (2) | 80.0% (8) | 10 |
| **HIV Viral suppression** |  |  |  |
| Suppressed | 15.8% (52) | 84.2% (278) | 330 |
| Non-suppressed | 54.6% (6) | 45.4% (5) | 11 |
| **Marital status** |  |  |  |
| Single | 31.7% (19) | 68.3% (41) | 60 |
| Married/cohabiting | 9.5% (19) | 90.5% (180) | 199 |
| Divorced/separated | 24.3% (17) | 75.7% (53) | 70 |
| widowed | 25% (3) | 75% (9) | 12 |

### Description of In-depth Interview participant characteristics

In total, 15 participants—eight women and seven men—who had not finished their IPT took part in the qualitative study. Four of the individuals were single, six were married or cohabitating, and five were divorced or legally separated. The median age was 34 overall (IQR: 28.5-45). One respondent lived in Mukono district, seven respondents lived in Kampala district, six respondents lived in Wakiso district. Four participants had completed their secondary education, four had completed their primary education, and one had completed their tertiary or university education (Table 3).

**Table 5:** Characteristics of IDI respondents.

| Characteristic | Number |
| --- | --- |
| **Sex** |  |
| Male | 7 |
| Female | 8 |
| **Age** |  |
| Overall median age | 34 |
| Median age (male) | 34 |
| Median age (female) | 35.5 |
| **Marital status** |  |
| Single | 4 |
| Married/cohabiting | 6 |
| Separated/divorced | 5 |
| **Education level** |  |
| Primary education | 14 |
| Secondary education | 4 |
| Tertiary or university | 1 |
| **District of Residence** |  |
| Kampala | 8 |
| Wakiso | 6 |
| Mukono | 1 |

### IPT completion

Of the 341 persons who started INH, 83% (283) patients finished the dose, compared to 17% (58) who did not. 86.4% (204) of the female participants in the IPT finished it, whereas 13.6% (32) did not. In terms of guys, 75.2% (79) finished IPT while 24.8% (26) did not. 84.2% (278) of patients with a suppressed HIV viral load finished IPT, while 15.8% (52) did not. 54.6% (6) of individuals with a non-suppressed HIV viral load did not finish IPT, compared to 45.4% (5) who did. IPT was successfully completed by 79.8% (183) of patients who were under the age of 29, compared to 20.2% (21) who were not. About 91% (180) of the patients who were married or cohabitated completed IPT, compared to 9.5% (19) who did not.

### Factors associated with IPT non-completion

#### Bivariable analysis

The prevalence of IPT non-completion was 23% lower among patients aged 30 years and above compared to those aged 29 years and below [Crude PR 0.77, CI: 0.48-1.25]. Males had an 83% higher prevalence of IPT non-completion compared to females [Crude PR 1.83, 95% CI: 1.15-2.91]. Patients with a non-suppressed HIV viral load had a 2.46 higher prevalence of IPT non-completion compared to those with a suppressed HIV viral load [Crude PR 3.46, 95% CI: 1.91-6.28]. Patients who were married/cohabiting had a 70% lower prevalence of IPT non-completion compared to those who were single [Crude PR 0.30, 95% CI: 0.17-0.53].

#### Multivariable analysis

In multivariable analysis, factors significantly associated with IPT non-completion were; sex, HIV viral suppression, and marital status. The prevalence of IPT non-completion among males was 2.24 times the prevalence among females [aPR 2.24, 95% CI: 1.40-3.58]. The prevalence of IPT non-completion among patients with a non-suppressed HIV viral load was 3.00 times the prevalence among those with a suppressed HIV viral load [aPR 3.00, 95% CI: 1.435-6.65]. Patients who were married/cohabiting had a 69% lower prevalence of IPT non-completion compared to those who were single [aPR 0.31, 95% CI: 0.17-0.55] (Table 6).

**Table 7:** Bivariable and multivariable analysis.

| Variable | Completed IPT<br>No % (n) | Yes % (n) | Crude<br>Prevalence<br>Ratios | Adjusted<br>Prevalence<br>Ratios (aPR) |
| --- | --- | --- | --- | --- |
| **Overall** | **17% (58)** | **83% (283)** | **N/A** | **N/A** |
| **Age** |  |  |  |  |
| 0-29 | 20.2%<br>(21) | 79.8% (83) | 1.00 | -- |
| 30+ | 15.6%<br>(37) | 84.4%<br>(200) | 0.77 (0.48-1.25) | -- |
| **Sex** |  |  |  |  |
| Female | 13.6%<br>(32) | 86.4%<br>(204) | 1.00 | 1.00 |
| Male | 24.8%<br>(26) | 75.2% (79) | 1.83 (1.15-2.91) * | 2.24 (1.40-3.58)<br>** |
| **WHO HIV clinical**<br>**stage** |  |  |  |  |
| Stage 1 | 16.9%<br>(56) | 83.1%<br>(275) | 1.00 | -- |
| Stage 2&3 | 20.0% (2) | 80.0% (8) | 1.18 (0.33-4.19) | -- |
| **HIV Viral load status** |  |  |  |  |
| Suppressed | 15.8 (52) | 84.2 (278) | 1.00 | 1.00 |
| Non-suppressed | 54.6 (6) | 45.4 (5) | 3.46 (1.91-6.28) *** | 3.00 (1.35-6.65)<br>** |
| **Marital status** |  |  |  |  |
| Single | 31.7 (19) | 68.3 (41) | 1.00 | 1.00 |
| Divorced/separated | 24.3 (17) | 75.7 (53) | 0.77 (0.44-1.34) | 0.77 (0.43-1.38) |
| Married/cohabiting | 9.5 (19) | 90.5 (180) | 0.30 (0.17-0.53) *** | 0.31(0.17-0.55)<br>*** |
| Widowed | 25 (3) | 75 (9) | 0.79 (0.28-2.26) | 1.07 (0.35-3.24) |
| **BMI** |  |  |  |  |
| Normal weight | 19.9%<br>(37) | 80.1%<br>(149) | 1.00 | -- |
| Underweight | 22.2% (4) | 77.8% (14) | 1.12 (0.45-2.78) | -- |
| Pre-obesity | 14.1% (12) | 85.9% (73) | 0.71 (0.40-1.30) | -- |
| Class 1 obesity | 12.1% (4) | 87.9% (29) | 0.61 (0.23-1.60) | -- |
| Class 2&3 obesity | 5.3% (1) | 94.7% (18) | 0.26 (0.38-1.83) | -- |

**Level of significance <0.05\*; <0.01\*\*; <0.001\*\*\***

#### Barriers to IPT completion

Themes and subthemes that emerged were;

Theme; Knowledge about IPT.

Theme: Challenges encountered while taking IPT. Subthemes: Lack of transport, Distance to the health facility, Pill burden, Relocation.

##### Theme; Knowledge about IPT

One of the reasons for non-compliance was the absence of IPT-related health education. The respondent was unaware of his IPT initiation.

> *“…I met a friend at the garage where I work who disclosed that he was on drugs and had not received IPT all his life. So, I wondered why I had been given these tablets yet the doctor didn’t tell me I had TB, I swallowed them for 2 months and got tired and stopped*” single male patient

##### Theme: Challenges encountered while taking IPT

###### Subtheme: High pill burden

High pill burden was another reason for IPT non-completion. This was more pronounced among patients that had to take other drugs for other pre-existing conditions such as hypertension in addition to ART and IPT.

> “…*the tablets to swallow are very many, I have these ART tablets, then my hypertension tablets, and then the IPT. So, at times I’d forget to swallow the IPT as it wasn’t treating me for anything and I completely stopped after forgetting it thrice*” Female Patient.

###### Subtheme: Distance to the health facility

Distance to the health facility was cited as a challenge as some of the patients could not afford transport every month to the health facility to pick up their IPT doses.

> “…*the first challenge, most of us come from far and at the beginning they first gave me for a month and most of my work I am casually employed. So sometimes it would be hard for me to tell my boss there is somewhere I am going I will not be around now*” Male Patient.

###### Subtheme: Relocation

Change of business and relocation from one place to another was cited as another reason for IPT non-completion

> “…*when I shifted from Kisenyi to Kitebi, I didn’t go with my card and when I went to get my drugs from Kitebi, they didn’t give me IPT yet in Kisenyi they had given me for three months. I explained to the doctors but they told me they’d give me next time until I got fed up and returned to Kisenyi where they have again started me on IPT again*” Female patient.

## Discussion

### IPT Completion

The goal of the study was to ascertain the IPT completion rate and related variables among those receiving antiretroviral therapy at Kisenyi Health Center IV in Kampala. The cohort of patients under study had an IPT completion rate of 83%, which was considered to be a comparatively high rate. This result is comparable to research from Zimbabwe and India, where studies (26, 27) indicated that 81% of HIV patients have completed IPT. This completion rate is higher than that of the SEARCH HIV test and treat trial, which was done in five communities in Uganda (28), and which Tram et al. reported at 73%.

The higher completion level found by our study can be attributed to the efforts of the national Quality Improvement (QI) team at the Ministry of Health founded in January 2019 to improve IPT completion as part of a wider continuous quality improvement effort in Uganda.

Situational analysis of IPT completion in high-volume facilities and coaching support for these locations across all regions 29 were part of the QI initiatives. The majority of patients receiving INH concurrently with ART lowers the expense of obtaining medication because patients wouldn’t need to make as many journeys to the medical institution, which contributes to the relatively high completion rate. Despite the fact that this study indicated that IPT completion rates have increased over time, 17% of patients who begin IPT do not finish the course of treatment. If IPT is not completed, its effectiveness is reduced and the patient is once again at danger of acquiring active TB, which raises morbidity and mortality.

### Factors associated with IPT non-completion among patients on ART

Males were shown to have a higher prevalence of IPT non-completion than females. This finding is consistent with a study conducted in rural Malawi by Little et al., which found that males were more likely than females to fail to complete the study (29). The typical masculine conduct, lower uptake of facility-based services, cultural precepts, and societal conventions that obstruct men’s health-seeking activities and lead to subpar treatment outcomes can all be used to explain this. To increase completion rates, this necessitates focused follow-up with male patients undergoing IPT.

The study also found that patients who had a non-suppressed HIV viral load had a higher prevalence of IPT non-completion. HIV viral non-suppression has been reported to be associated with; having active TB, HIV clinical stage, duration of ART, ART regimen, baseline CD4, and ART adherence among other factors (30-32). Additionally, HIV viral non-suppression has been reported to be associated with adherence; defined as a patient’s ability to follow a treatment plan, take medications at prescribed times and frequencies and follow restrictions regarding food and other medications (33). Patients who are HIV virally non-suppressed are more likely to have poor ART adherence (30, 34). This, therefore, explains the association between IPT non-completion and viral non-suppression among HIV patients.

Patients who did not complete IPT were more likely to be HIV virally non-suppressed. It is possible that non-virally suppressed patients were likely to be non-adherent to ART and other associated medicines including IPT. Targeted follow-up of HIV virally non-suppressing patients on IPT is needed to reduce the prevalence of IPT non-completion in this group.

Additionally, the study found that marriage was strongly associated with a lower rate of IPT non-completion than was being single. Married individuals are more likely to be open about their status with their partners (35), which offers emotional and psychological support (36), which has been linked to improved treatment outcomes (37, 38).

This can also be explained by the fact that marriage gives patients stability by allowing them to be taken care of while undergoing therapy. IPT completion rates are expected to increase if these patients’ wives provide them meals on time and remind them to take their medications.

This finding highlights the importance of providing ART patients, as well as those on IPT, with social support networks. These patients can benefit from social and psychological support from services like psychosocial and adherence counseling as part of the HIV care program (39).

### Reasons for IPT non-completion

One of the causes of IPT non-compliance, according to this study, is a lack of health education on the topic among HIV patients. In order to improve health literacy, encompassing knowledge and life skills that support individual and communal health, health education entails consciously creating learning opportunities requiring some sort of communication (40). Numerous studies have shown that health education can enhance the effectiveness of patient care (41, 42).

The lack of health education among HIV patients on the other hand is related to lower patient adherence and poor treatment outcomes (43, 44). Due to the big volumes of patients that the health workers attend to on clinic days (23, 45), health workers tend to focus on providing drug refills neglecting to provide health education (46). In these circumstances, patients receive drug refills without health education on the benefits of the drugs they have received. These patients are more likely not to adhere to treatment resulting in non-completion.

Some of the respondents abandoned IPT due to the pill burden, especially for those that were on medication for other underlying health conditions such as diabetes and hypertension. This finding is similar to studies conducted in Kenya, Swaziland, Zimbabwe, and India which found that pill burden was one of the reasons for IPT non-completion, especially for patients that had to take other drugs for underlying conditions such as hypertension in addition to ART (27, 47-50). This can be explained by the difficulty in concurrent management of all conditions as it can be complicated not only by high pill burden and increased risks of drug-drug interactions but also by overlapping toxicities and immune reconstitution inflammatory syndrome (51).

IPT regimens with less frequent dosages compared to INH have been shown to be as effective for TB prevention as INH but with better safety and completion levels. A 3-month weekly course of rifapentine and Isoniazid (3-HP) is as effective for TB prevention as INH, but with better safety and completion levels (52). Other regimens including 4-months of daily Rifampicin (4R), and 3-4 months of daily Isoniazid and Rifampicin (3-4HR) have also been shown to be effective for TB prevention, safe, and for shorter durations compared to 6-9-month course of INH (53-55). Regimens with shorter durations and less frequent dosages should be considered for patients with pill burden challenges to improve IPT completion levels.

Distance to the health facility was cited as one of the barriers to IPT completion as patients could not afford transport every month to the health facility to pick up their IPT doses. This echoes findings from studies conducted in Tanzania, Zimbabwe, and Ethiopia which found that patients were discouraged by the need to attend clinic every month combined with the long treatment duration of IPT (26, 56, 57). A significant proportion of patients do not get ART from their nearest facility (58, 59) due to various reasons that include the fear of stigma, and the perception that care at local facilities being of lower quality compared to distant ones (59, 60).

This additional distance that the patient has to travel is compounded by unpredicted commodity shortages resulting from challenges in the supply chain management (61) which leads the health workers into giving IPT doses for fewer months compared to ART that was given for a longer period. This results in the need for the patient to attend the ART clinic more frequently to obtain refills which translates into additional costs on the patient’s side but also crowding at the clinic.

Uganda in 2016 adopted Differentiated Service Delivery (DSD) models for both HIV testing services and treatment to cater to the needs and preferences of the increasing number of clients as well as improve the quality of services and efficiency in service delivery (23). Community-based DSD models have been documented to have benefits both to the patient and the health facility in terms of reducing the cost of obtaining ART on the patient’s side, improved patient management, and reduced patient congestion at the health facility (62). These DSD models can further be utilized for giving IPT refills to patients in addition to ART.

Another barrier to IPT completion among HIV patients was relocation from one place to another. When patients relocate to new areas within or outside of Kampala, they also transfer to nearby health facilities. The majority of these patients transfer to these sites without proper referral documentation in what is termed “self-transfer” (63). In this case, the transfer-in facility may not offer care continuity (in this case INH) due to a lack of relevant documentation from the transfer-out facility (64). This finding is similar to a study done in Dar es salaam where people living with HIV who were transferred to other clinics within Dar es Salaam region after IPT initiation had significantly lower IPT completion levels compared to those that attended the same clinic (57). These findings are also similar to studies conducted in Zimbabwe and Ethiopia (26, 65). This can also be explained by patients’ failure to utilize health facilities in their new localities (63). Linking patient databases to improve access and continuity of care can be explored to improve not only IPT completion but also other treatment like ART.

### Study limitations

The study limitations were mainly related to missing and incomplete data since this was a retrospective record review of routinely collected program data. This was overcome by triangulating data from different sources i.e., IPT, ART registers, ART cards, and the UgandaEMR.

The study was conducted in one health facility in Kampala city and hence the findings may not be generalizable to other health facilities within other cities and districts in Uganda. Treatment completion was assessed by a patient picking up a refill as a proxy for actual consumption of isoniazid. It is however possible that patients might have returned to pick up another bottle of tablets without consuming all the tablets from the previous refill.

The study also explored barriers to IPT completion from the patient’s perspective and did not look at the provider and system-related barriers to IPT completion. To effectively improve IPT use, barriers to its completion from the provider’s perspective need to be explored as well.

Some of the planned interviews by stratification were not conducted as there were no patients/respondents in those strata. The barriers to IPT completion presented represent the patient categories that were interviewed.

The barriers to IPT completion explored may have been limited by social desirability bias. This was mitigated by employing unconditional positive regard (66).

### Recommendations

1. The health caregivers/health workers need to provide patients with IPT-related health education before IPT initiation. Opportunities to give group health education on clinic days can be leveraged. Patient attitudes towards the therapy and consequently completion levels are likely to improve when patients are fully aware of why they are being initiated on the treatment.
2. There’s a need for the Ministry of Health and partners to increase availability and access to other regimens of IPT with less frequent dosages compared to a 6-9 months daily dose of INH. This will reduce the pill burden, especially on patients with comorbidities, improve adherence and completion levels.
3. Health workers and health caregivers should provide and intensify treatment adherence support and counseling to HIV patients who are virally non-suppressed, not only for ART but also IPT.
4. There’s a need for the Ministry of health to explore the use of biometric systems and linking ART/TB clinics across health facilities to enable patients especially the mobile populations to access ART and IPT at any accredited health facility but also with accurate and timely documentation.
5. Health facilities should take advantage of community DSD models and exploit them to deliver IPT in addition to ART and other related medications to eligible patients. Additionally, multi-month ART refills synchronized with IPT should also be taken advantage of to improve uptake and completion of IPT.
6. There’s need for further research on factors influencing IPT completion from the health system and provider’s perspective.

## Conclusion

The IPT completion level was found to be relatively high, at 83% among the cohort of patients but still, less than the expected 95%. This corresponds with a general improvement in HIV services and the monitoring of HIV services by health facilities and stakeholders. Being a male, and being HIV virally non-suppressed were significantly associated with a higher prevalence of IPT non-completion. Being married was associated with a lower prevalence of IPT non-completion. Lack of health education, pill burden, distance to the health facility, and relocation were reported as the reasons for failure to complete IPT.

## Data Availability

The data underlying the results presented in the study are available from https://drive.google.com/file/d/1-VCy-JfMOuDt1d24e7kOdO_iBjHNWY-2/view?usp=sharing

## Acknowledgments

Special thanks go to Vicky Kyobutungi for providing a second pair of eyes, and for the input to this work. Thanks also go to Muzoora Enock, Mirembe Annah and Namara Rebecca for the moral support accorded throughout this work.

I am also grateful to the Directorate of Public health and Environment at KCCA, administrators, and health workers of Kisenyi Health Centre IV who provided all the records and data that were needed for this research. Special thanks go to all respondents who took part in this study.

I also acknowledge the support accorded by my colleagues; Tusabe Joan, Tabwenda Lilian, Nanfuka Harriet, Kisenyi HC IV staff; Thadoues Mugenyi, and Gerald Mwesigye.

## Author Contributions

**Conceptualization:** Ian Amanya, Anthony Ssebagereka, Richard Mugambe.

**Formal analysis:** Ian Amanya, Michael Muhoozi.

**Funding acquisition:** Ian Amanya.

**Investigation:** Ian Amanya, Anthony Ssebagereka, Richard Mugambe.

**Methodology:** Ian Amanya, Anthony Ssebagereka, Richard Mugambe.

**Project administration:** Ian Amanya.

**Resources:** Ian Amanya.

**Software:** Ian Amanya, Michael Muhoozi.

**Supervision:** Ian Amanya, Anthony Ssebagereka, Richard Mugambe.

**Visualization:** Ian Amanya.

**Writing – original draft:** Ian Amanya, Dickson Aruhomukama, Anthony Ssebagereka, Richard Mugambe.

**Writing – review & editing:** Ian Amanya, Dickson Aruhomukama, Anthony Ssebagereka, Richard Mugambe.

